# Diffusion MRI Metrics and their Relation to Dementia Severity: Effects of Harmonization Approaches

**DOI:** 10.1101/2021.10.04.21263994

**Authors:** Sophia I. Thomopoulos, Talia M. Nir, Julio E. Villalon-Reina, Artemis Zavaliangos-Petropulu, Piyush Maiti, Hong Zheng, Elnaz Nourollahimoghadam, Neda Jahanshad, Paul M. Thompson, for the Alzheimer’s Disease Neuroimaging Initiative

**Author notes:** Data used in preparing this article were obtained from the Alzheimer’s Disease Neuroimaging Initiative (ADNI) database (adni.loni.usc.edu). As such, many investigators within the ADNI contributed to the design and implementation of ADNI and/or provided data but did not participate in analysis or writing of this report. A complete listing of ADNI investigators can be found at: http://adni.loni.usc.edu/wpcontent/uploads/how_to_apply/ADNI_Acknowledgement_List.pdf.

## Abstract

Diffusion-weighted magnetic resonance imaging (dMRI) is sensitive to microstructural changes in the brain that occur with normal aging and Alzheimer’s disease (AD). There is much interest in which dMRI measures are most strongly correlated with clinical measures of AD severity, such as the clinical dementia rating (CDR), and biological processes that may be disrupted in AD, such as brain amyloid load measured using PET. Of these processes, some can be targeted using novel drugs. Since 2016, the Alzheimer’s Disease Neuroimaging Initiative (ADNI) has collected dMRI data from three scanner manufacturers across 58 sites using 7 different protocols that vary in angular resolution, scan duration, and in the number and distribution of diffusion-weighted gradients. Here, we assessed dMRI data from 730 of those individuals (447 cognitively normal controls, 214 with mild cognitive impairment, 69 with dementia; age: 74.1±7.9 years; 381 female/349 male). To harmonize data from different protocols, we applied ComBat, ComBat-GAM, and CovBat to dMRI metrics from 28 white matter regions of interest. We ranked all dMRI metrics in order of the strength of clinically relevant associations, and assessed how this depended on the harmonization methods employed. dMRI metrics were associated with age and clinical impairment, but also with amyloid positivity. All harmonization methods gave comparable results while enabling data integration across multiple scanners and protocols.

## 1. INTRODUCTION

Alzheimer’s disease (AD) is the most prevalent form of dementia, affecting over 50 million people worldwide – a number which is expected to grow to 82 million in the next decade.^1^ Brain imaging techniques such as magnetic resonance imaging (MRI) and positron emission tomography (PET) are now widely used to track and monitor AD progression; with the recent approval of anti-amyloid treatments for AD, there is a great need to better understand risk factors that influence the component biological processes of brain aging and AD, and to screen people for enrolment in clinical trials. Although the definition of AD has recently evolved to include measures of amyloid and tau proteins, assessed using PET or lumbar puncture to assess CSF,^2^ the high invasiveness, cost, and limited availability of these procedures has stimulated interest in other imaging measures that track AD progression, specifically variants of MRI, such as diffusion MRI, ASL, and resting state functional MRI. Even so, the relation between neurodegeneration and microstructural brain differences on MRI to brain amyloid load and clinical measures of disease severity is not fully understood.

Biomarkers help to understand and track biological processes involved in brain aging and AD by providing proxy measures of relevant pathophysiology.^3^ Among the many subtypes and variants of MRI used in AD research, diffusion-weighted MRI (dMRI) provides insight into microstructural brain changes that occur with aging and dementia, especially in the white matter (WM), that are not detectable with standard anatomical MRI. Diffusion tensor imaging (DTI) has been used as an overall *in vivo* proxy measure for WM tissue integrity, which also declines with normal aging.^4^ AD progression is associated with neuronal and glial cell loss, loss of long and short range axonal connections, and myelin degeneration. DTI-derived metrics, including fractional anisotropy (FA), mean (MD), radial (RD) and axial diffusivity (AxD), are frequently used to quantify WM microstructure. Recent innovations with multi-shell diffusion protocols have also been developed to model and assess intra- and extracellular diffusion, neurite dispersion, and diffusion kurtosis (e.g., NODDI^5^, MAP-MRI^6^, and DKI^7^).

The Alzheimer’s Disease Neuroimaging Initiative (ADNI) is a large-scale, longitudinal, multisite initiative that evaluates imaging, clinical and molecular biomarkers of AD;^8^ the imaging component includes various MRI modalities, such as diffusion MRI. In response to the criticism that studies with small sample sizes may lead to non-reproducible effects,^9^ there has been a shift in neuroimaging research towards pooling datasets, increasing sample sizes, and including historically excluded populations.^10^ Since its launch in 2004, ADNI has become a valuable public dataset that has been widely analyzed in hundreds of publications.^11^ In its second phase (ADNI2), a diffusion imaging protocol was added at around one third of acquisition sites, all of which used General Electric (GE) scanners. To increase the number of dMRI scans made available, additional vendor-specific dMRI protocols were implemented for GE, Siemens and Philips scanners during ADNI’s third phase (ADNI3).^12^ Consequently, in addition to site, a new potential “confound” was introduced into the study’s dMRI data – that of scanner manufacturer and acquisition protocol.

Determining the best way to harmonize multisite data is now a prime focus in neuroimaging.^13^ In prior studies,^14–16^ we showed that while it is possible to combine dMRI data from multiple centers using a random effects design to model effects of protocol, there is growing interest in novel ways to mitigate site- and protocol-related effects in multisite research.^16^ Harmonized preprocessing protocols may help to reduce some variability,^17–19^ but multisite studies such as ADNI are affected by ‘site’ or ‘batch’ effects, including but not limited to: (1) technical factors (e.g., acquisition site, scanner manufacture, scanner model and coil), and (2) cohort factors, i.e., differences or selection biases in the individuals assessed at each site.

### 1.1 Relevant Prior Work

Since our previous work,^16^ there have been numerous developments in the field of MRI harmonization.^20,21^ Some methods harmonize image-derived data at the statistical level, for example, by modeling site as a random effect. Other methods adjust the raw scan data, using generative adversarial methods such as CycleGANs or StarGANs that adjust scans to appear as if they were collected using a different scanner or protocol. Moyer et al.^22^ harmonized multisite diffusion MRI data by using a variational autoencoder to compute a site-invariant embedding of the raw data. They then reconstructed data to match that collected at any site by combining the latent space vector with a site-dependent code. Dinsdale and colleagues^23^ showed that such a site-invariant embedding can boost task performance on machine learning tasks that use multisite data, such as brainAGE estimation. Liu et al.^24^ used a StarGAN method to separate the ‘content’ and ‘style’ of a set of images, to interconvert T1-weighted MRI data across sites and scanners. Zuo et al.^25^ recently developed CALAMITI, which uses multimodal data from the same individual to learn the manifold of different MR contrasts. By disentangling participant and site effects, the authors adjusted individual data to match any given scanning protocol. Direct GAN-based adjustments of scans are under intense investigation; even so, approaches that statistically adjust measures derived from images are currently more widely used, as they can also be applied when the raw scan data cannot readily be shared due to privacy constraints, or when the amount of data may be insufficient to train a GAN to produce stable corrections.

ComBat, which was originally proposed by Johnson et al.^26^ as a batch adjustment method for genomics data, falls into the latter category. ComBat uses an empirical Bayes framework to fit a set of covariates to the data, and applies a linear shift-and-scale transform to the residuals of the model to match a reference distribution, before adding back the covariate effects to yield an adjusted set of data for each site. Variations stemming from the original ComBat have been developed for neuroimaging data, three of which are assessed here: (1) ComBat was initially adapted for neuroimaging data by Fortin et al.^27^, otherwise referred to as neuroComBat; (2) to model age trends more accurately, Pomponio et al.^28^ proposed ComBat-GAM, which uses a generalized additive model (GAM); (3) CovBat, proposed by Chen at al.^29^, aims to remove site or scanner effects in the mean, variance, and spatial covariance of the derived measures. These methods try to regress out effects induced by site; other approaches such as hierarchical Bayesian regression model, each site’s data distribution using a hyperprior, which models the distribution of the site means and variances.^30,31^

While we previously applied ComBat to the dMRI metrics available from an initial ADNI3 dataset^16^ which was 43% of the current sample size, more recent methods such as ComBat-GAM and CovBat have not yet been applied to ADNI3 dMRI indices, to the best of our knowledge. Here we extend our work^16^ by applying a new dMRI preprocessing protocol on the updated cohort and exploring the harmonization capabilities, at an ROI level, of ComBat, ComBat-GAM and CovBat which all aim to remove site-effects but preserve clinically relevant between-subject biological variability. Our goal was (1) to understand how harmonization choices affect associations between dMRI and clinically relevant measures of disease burden in dementia, and (2) to top up the sample from our prior reports, to include the much larger sample now available.

## 2. METHODS

### 2.1 Participants

Baseline ADNI3 demographic, cognitive and neuroimaging data analyzed in this article were obtained from the ADNI database (ida.loni.usc.edu). Subjects who were missing basic clinical information or had poor-quality imaging data, such as scans with severe motion, distortion, or ringing, were not included in the final sample of N=730. Clinical scores known to characterize AD, specifically the Clinical Dementia Rating sum-of-boxes (CDR-sob)^32^, and amyloid beta (Aβ) status were obtained where available. Aβ-status, i.e., positive (Aβ+) or negative (Aβ–), was determined by either mean 18F-florbetapir (Aβ+ defined as >1.11)^33,34^ or florbetaben (Aβ+ defined as >1.20)^35,36^ PET cortical SUVR uptake, normalized by using a whole cerebellum reference region. The overall sample included 447 cognitively normal controls (CN; 185 male / 262 female; 55.1–95.3 years old; mean age: 73.2±0.75 years; 115 Aβ+ / 225 Aβ-; 0.07±0.27 CDR-sob), 214 participants with mild cognitive impairment (MCI; 123 male / 91 female; 55.1–93.2 years old; mean age: 75.1±8.1 years; 79 Aβ+ / 87 Aβ-; 1.39±1.10 CDR-sob) and 69 with dementia (41 male / 28 female; 55.2–96 years old; mean age: 77.4±8.9 years; 51 Aβ+ / 9 Aβ-; 5.45±2.77 CDR-sob). **Table 1** summarizes participant demographic information and cognitive measures, stratified by acquisition protocol.

**Table 1.** Participant demographics. ‘Site N’ denotes the number of sites across North America using the dMRI acquisition protocol specified (protocols are further detailed in **Table 2**).

| Protocol | Total N | Diagnosis | | | M | F | Age<br>(Mean±SD) | CDR-sob<br>(Mean±SD) | Amyloid | | Site N |
| --- | --- | --- | --- | --- | --- | --- | --- | --- | --- | --- | --- |
| | | CN | MCI | Dementia | | | | | Aβ+ | Aβ- | |
| **S55** | 270 | 175 | 71 | 24 | 120 | 150 | 74.4±7.9 | 0.93±2.0 | 103 | 117 | 18 |
| **S31** | 96 | 59 | 27 | 10 | 39 | 57 | 72.0±8.5 | 0.96±1.8 | 26 | 49 | 10 |
| **S127** | 112 | 68 | 36 | 8 | 53 | 59 | 73.7±7.6 | 0.78±1.5 | 35 | 58 | 14 |
| **P36** | 46 | 20 | 23 | 3 | 24 | 22 | 73.1±6.9 | 0.84±1.1 | 12 | 27 | 5 |
| **P33** | 56 | 37 | 14 | 5 | 34 | 22 | 75.7±7.7 | 1.03±2.1 | 19 | 21 | 6 |
| **GE36** | 42 | 21 | 17 | 4 | 24 | 18 | 72.2±7.1 | 1.29±2.7 | 13 | 22 | 4 |
| **GE54** | 108 | 67 | 26 | 15 | 55 | 53 | 76.1±8.1 | 1.16±2.0 | 37 | 57 | 10 |
| **Total** | **730** | **447** | **214** | **69** | **349** | **381** | **74.1±7.9** | **0.97±1.9** | **245** | **351** | **58** |

### 2.2 MRI Data Acquisition

Acquisition parameters for available ADNI3 dMRI data (as of December 2020) are shown in **Table 2**. All dMRI were acquired with an isotropic 2×2×2 mm^3^ voxel size. Diffusion-weighted and anatomical T1-weighted (T1w) MRI MP-RAGE data (independent of scanner vendor and model: isotropic voxel size of 1×1×1mm^3^, 208×240×256mm^3^ matrix, TE= 2.98 ms, TR=2300 ms, TI=900 ms, 11° flip angle, scan duration: 6 mins 20 secs) were acquired on 3.0 Tesla scanners from Siemens, Philips or GE sites across 58 centers in North America. More information on the ADNI MRI protocols may be found here: http://adni.loni.usc.edu/methods/documents/mri-protocols/.

**Table 2.** ADNI3 dMRI acquisition protocols.

| Protocol | Scanner |  | Volumes |  |  | Matrix | Acquisition Time (min:sec) |
| --- | --- | --- | --- | --- | --- | --- | --- |
|  | Vendor | Model | b<sub>0</sub> | DWI | Total |  |  |
| **GE36** | GE | Basic Widebore 25x | 4 b=0 s/mm<sup>2</sup> | 32 b=1000 s/mm<sup>2</sup> | 36 | 256x256 | 9:52 |
| **GE54** | GE | Basic 25x | 6 b=0 s/mm<sup>2</sup> | 48 b=1000 s/mm<sup>2</sup> | 54 |  | 7:09 |
| **P33** | Philips | Basic Widebore | 1 b=0 s/mm<sup>2</sup> | 32 b=1000 s/mm<sup>2</sup> | 33 | 128x128 | 7:32 |
| **P36** | Philips | Basic Widebore R3 | 1 b=0 s/mm<sup>2</sup><br>3 b=2 s/mm<sup>2</sup> | 32 b=1000 s/mm<sup>2</sup> | 36 |  | 6:54 |
| **S31** | Siemens | Basic VB17 | 1 b=0 s/mm<sup>2</sup> | 30 b=1000 s/mm<sup>2</sup> | 31 | 116x116 | 7:02 |
| **S55** | Siemens | Basic Skyra E11 & Prisma D13 | 7 b=0 s/mm<sup>2</sup> | 48 b=1000 s/mm<sup>2</sup> | 55 |  | 9:18 |
| **S127** | Siemens | Advanced Prisma VE11C | 13 b=0 s/mm<sup>2</sup> | 6 b=500 s/mm<sup>2</sup><br>48 b=1000 s/mm<sup>2</sup><br>60 b=2000 s/mm<sup>2</sup> | 127 |  | 7:25* |

### 2.3 MRI Data Preprocessing

Building on the preprocessing protocol previously developed by the ADNI Diffusion MRI Core, as described in Nir at al.^14^, we updated our preprocessing pipeline (**Figure 1**) after assessing^37^ new tools that have now become available and may reduce common sources of noise and artifacts and improve model fit.

**Figure 1.**
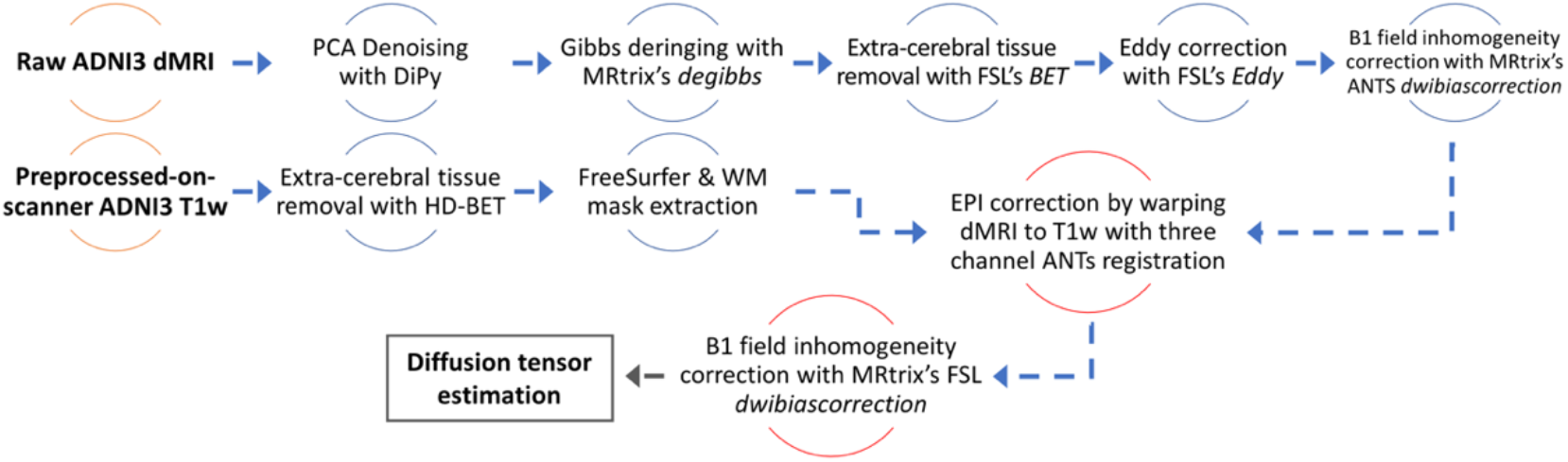
MRI preprocessing pipeline flowchart.

Raw dMRI were first denoised using DiPy’s^38^ principal component analysis (PCA)^39^ denoising algorithms. For data that were zero-padded in *k*-space (i.e., GE) noise was estimated using a local PCA (LPCA)^39^ algorithm. For data with an original acquisition matrix (i.e., Siemens and Philips) noise was estimated using the Marchenko-Pastur Estimation based denoising (MPPCA)^40,41^. Gibbs artifacts^37^ were corrected for using MRtrix’s^42^ *degibbs*^43^, and extracerebral tissue was removed using FSL’s^44–46^ *BET*. Eddy correction was performed using FSL’s *eddy_cuda*^47^ tool with *repol* outlier^48^ and “slice-to-volume” correction^49^, and bias field inhomogeneity was corrected with MRtrix’s *dwibiascorrection* ANTS function. To match the single-shell data, shells with b=500 and b=2000 s/mm^2^ were at this stage removed from the multi-shell protocol (i.e., S127) dMRI thus leaving the scans with 13 b=0 and 48 b=1000 shells. All metrics assessed were derived from b_0_ and b=1000 shells.

ADNI3 dMRI data is not acquired with reversed phase-encode blips, thus the *eddy_cuda* top-up tool could not be used. To compensate for this, we used registrations to T1w anatomical images to further correct for echo-planar imaging (EPI) distortions in the diffusion images. All ADNI3 T1w images available on the ADNI database were initially preprocessed – including gradient warp and intensity inhomogeneity correction – by vendor product on the scanner at acquisition time. The ADNI-preprocessed T1w data were skull stripped using HD-BET^50^, and run through FreeSurfer’s^51^ standardized pipeline (version 5.3) for further processing.

The subject-level preprocessed average B_0_ image was linearly aligned to the subject’s respective T1w using boundary-based registration (BBR)^52^ with the WM mask extracted with FreeSurfer. This output matrix was then applied to the preprocessed dMRI volumes. A three-channel ANTS^53,54^ non-linear registration, driven equally by B_0_, DTI and MD, and FA images, was used to warp dMRI to the T1w image. The linear and non-linear registrations were concatenated and inverted and applied to unregistered dMRI to both EPI correct images and bring them back to their native space with only one interpolation. The EPI-corrected DWI were then run through FSL’s bias field inhomogeneity correction using MRtrix’s *dwibiascorrection*. All outputs were visually inspected before inclusion in our final dataset, including any masks which were further manually edited when needed.

### 2.4 White Matter Tract Atlas ROI Summary DTI Metrics

Standard diffusion tensor derived metrics – FA, AxD, MD, RD – were computed using DiPy’s DTI weighted least squares function.

The JHU ICBM-DTI-81^55^ atlas FA was first warped to each subject’s FA using FSL’s *flirt* with 12 degrees of freedom followed by a nonlinear ANTS^56^ registration. The transformations were applied to stereotaxic JHU WM atlas labels^55^ (JHU MNI single subject WMPM1; **Table 3**) using nearest neighbor interpolation. A robust mean was calculated across voxels within each of 28 ROI using an iterative *M*-estimator from the ‘WRS2’ package in R.^57,58^

**Table 3.** Measures from 27 ROIs (averaged across left and right hemispheres) and the full WM were assessed.

|  |  |  |  |
| --- | --- | --- | --- |
| SCP | Superior cerebellar peduncle | CGH | Cingulum (hippocampus) |
| MCP | Middle cerebellar peduncle | Fx/ST | Fornix (*crus*) / *Stria terminalis* |
| ICP | Inferior cerebellar peduncle | SLF | Superior longitudinal fasciculus |
| PCT | Pontine crossing tract | SFO | Superior fronto-occipital fasciculus |
| ML | Medial lemniscus | SS | Sagittal *stratum* |
| CP | Cerebral peduncle | EC | External capsule |
| CST | Corticospinal tract | UNC | Uncinate fasciculus |
| ALIC | Anterior limb of internal capsule | PTR | Posterior thalamic radiation |
| PLIC | Posterior limb of internal capsule | TAP | *Tapetum* |
| RLIC | Retrolenticular part of internal capsule | Fx | Fornix |
| ACR | Anterior *corona radiata* | GCC | *Genu* of corpus callosum |
| SCR | Superior *corona radiata* | BCC | Body of corpus callosum |
| PCR | Posterior *corona radiata* | SCC | *Splenium* of corpus callosum |
| CGC | Cingulum (cingulate gyrus) | Full WM | Full white matter |

### 2.5 ROI Harmonization Methods using ComBat Variations

The three ComBat variations, including ComBat^27^, ComBat-GAM^28^ and CovBat^29^, were separately applied to the participants’ ROI-level FA, AxD, MD and RD data in order to harmonize derived measures across acquisition protocols (**Figure 2A**). Site-level adjustments using data harmonization methods were not attempted at this stage as many sites had N<10 subjects. Across all three ComBat variants, age, sex, age*sex and disease severity were designated as biological variables

**Figure 2.**
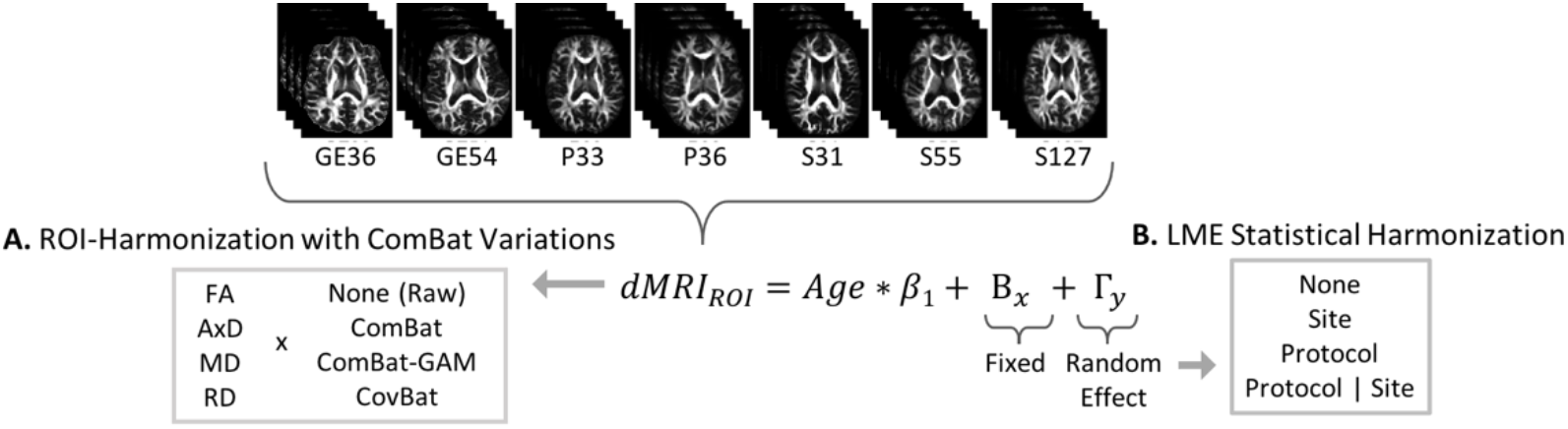
ROI and statistical harmonization method permutations applied to the data for assessment. Four ROI and four statistical approaches were evaluated, resulting in 16 total permutations.

#### 2.5.1 ComBat

ComBat was implemented using R scripts (https://github.com/Jfortin1/neuroCombat_Rpackage/) running on RStudio (ver. 1.4.1717) and R (ver. 3.6.1).

#### 2.5.2 ComBat-GAM

Quadratic effects of age were additionally included in the model. The ComBat-GAM approach was implemented using Python (ver. 3.8) scripts (https://github.com/rpomponio/neuroHarmonize).

#### 2.5.3 CovBat

A 95% parametric variance threshold was set for CovBat. CovBat was implemented using R scripts (https://github.com/andy1764/CovBat_Harmonization) running on RStudio (ver. 1.4.1717) and R (ver. 3.6.1).

### 2.6 Statistical Analyses

#### 2.6.1 Assessing Effects of Age, Cognitive Measures and Amyloid Positivity on dMRI Indices

Across all participants, the effects of age, CDR-sob and Aβ-status were assessed on the ‘raw’ (i.e., un-harmonized) and ComBat-harmonized ROI dMRI indices using multiple linear regressions. In all statistical models age, sex, age*sex, CDR-sob and Aβ-status were included as fixed effects. The effect of age was additionally assessed only in the subset of cognitively normal participants (N=447) using only sex and age*sex as fixed effects covariates. We corrected for multiple comparisons across ROIs using the false discovery rate procedure (FDR; *q*=0.05).^59^ For continuous variables, i.e., age and CDR-sob, effect sizes were calculated as *d-value=(2*t-value)/sqrt(degrees of freedom)*. Cohen’s *D* effect sizes are reported for dichotomous variables, i.e., Aβ-status.

#### 2.6.2 Statistical Harmonization Methods using Random Effects

In addition to harmonizing data at the ROI level, linear mixed-effects models (LME) were also used to model acquisition site and dMRI protocol at the statistical level using the *‘lme’* package in R. For the ‘raw’ ROIs and each of the three variations of ComBat-harmonized ROIs, in addition to the aforementioned fixed effects variables, three random effects grouping variables were used: (1) dMRI acquisition protocol, (2) acquisition site, or (3) site nested in dMRI protocol (protocol|site) (**Figure 2B**). For the remainder of the paper, we use the following notation to identify the harmonization permutation evaluated: *ROI-Harmonization Method* (*Random Variable*), where *ROI-Harmonization Method* is one of four methods identified in **Figure 2A** and *Random Variable* is one of four variables listed in **Figure 2B** (e.g., CovBat(None)).

## 3. RESULTS

### 3.1 ROI Harmonization Methods using ComBat Variations

Box plots of subject-level dMRI indices, split by dMRI protocol and diagnostic group, revealed that the three ComBat methods yielded largely similar results (**Figure 3**); the mean and variance of the average full WM MD and FA values were similar regardless of ROI-harmonization approach. In CN, the mean FA remained stable at 0.35 before and after harmonizing with each of the ComBat variations, but the range of values from 0.22–0.42 to 0.20–0.41. Respectively, in MCI, the mean FA remained stable at 0.34, and the range shifted from 0.32–0.42 to 0.26–0.41. In the dementia group, the mean FA remained stable at 0.33 and the range shifted from 0.22–0.42 to 0.25–0.39. As with FA, the mean full WM MD remained stable at 0.0008 and with a range of values between 0.0007–0.0009 before and after harmonizing with Combat and CovBat, but shifted to a mean of 0.0006 and range of 0.0005–0.0008 with ComBat-GAM. In MCI, we see similar results in the mean and range for full WM MD which remained stable at 0.008 and 0.007-0.009, but dropped to a 0.0006 mean and 0.0005–0.0008 range with ComBat-GAM. In the dementia group, the mean MD of full WM remained stable at 0.0008 but dropped to 0.0007 with ComBat-GAM. Pairwise *t*-tests between the four datasets (‘raw’, ComBat, ComBat-GAM, CovBat) did reveal significant differences (*p*<2.20×10^−16^) in all 28 ComBat-GAM harmonized MD and RD indices when compared to ‘raw’, ComBat or CovBat harmonized indices (**Figure 4**). FA was the most stable across comparisons, although 16 ROIs were significantly different when comparing ComBat and ComBat-GAM (*p*=0.003– 0.004).

**Figure 3.**
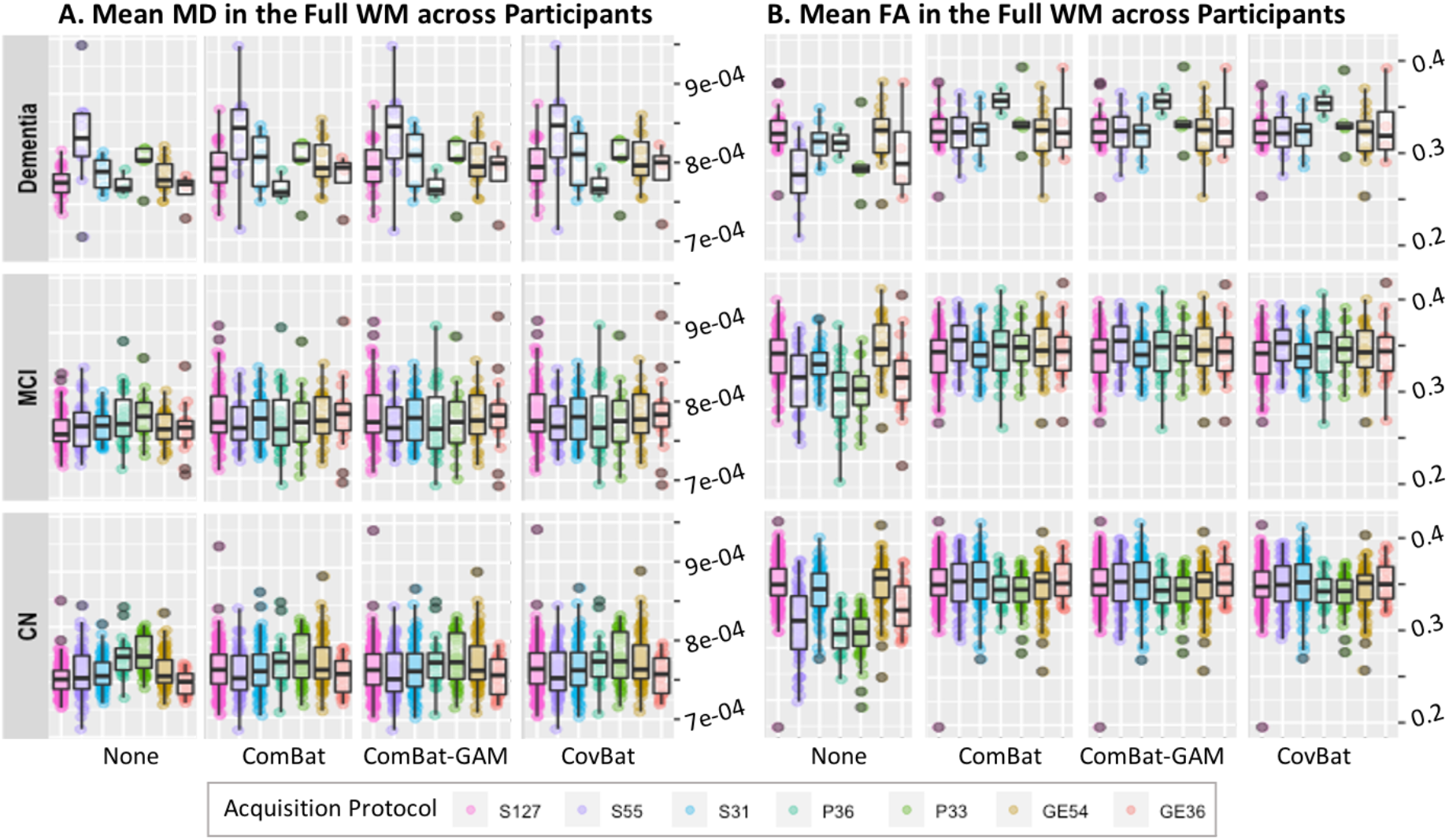
Scatter plot of subject-level average MD and FA in the full white matter, split by dMRI protocol, ComBat harmonization method, and diagnostic group. The effects of ComBat and its variants on the dMRI group means are apparent, especially for FA, and in MCI and CN participants, who make up the majority of the ADNI3 cohort (*right panels, last two rows*).

**Figure 4.**
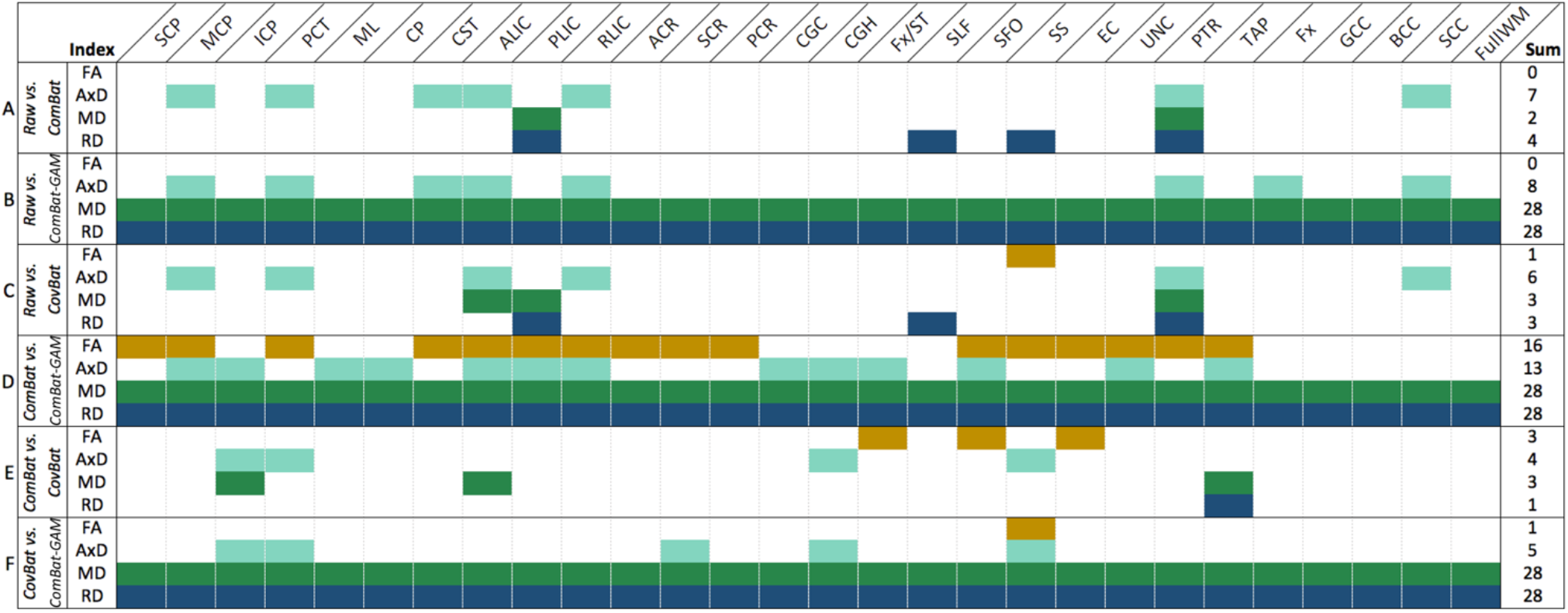
For each DTI measure, paired *t*-tests between ‘raw’ and harmonized dMRI ROIs across all subjects were run, with significant differences (*p*-values < 0.05) denoted in color. Each color represents a dMRI index: FA (gold), AxD (cyan), MD (dark green), RD (dark blue). The last column summarizes the total number of significant ROI differences for each comparison.

In CN participants, pairwise comparisons of dMRI indices from each protocol were compared before and after ComBat harmonization; protocol, age, sex, age*sex, Aβ-status and CDR-sob were modeled as fixed effects. While significant protocol differences were detected across pairwise protocol comparisons using the ‘raw’ dMRI ROIs, after harmonization using any of the three ComBat approaches, almost no protocol differences were detected (**Table 4**). The greatest number of ROI differences remained between the FA and RD derived from S127 and P33, regardless of ComBat variation.

**Table 4.**
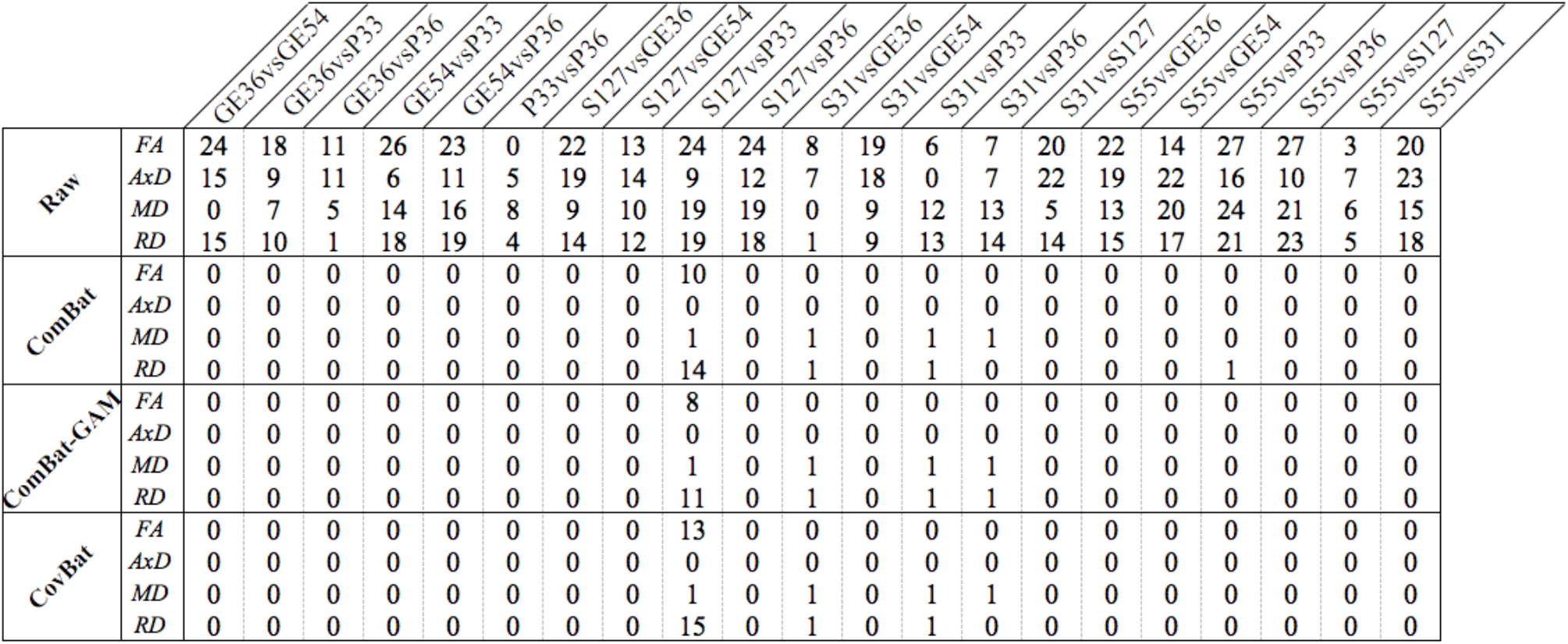
Total number of ROIs that were significantly different in pairwise comparisons of dMRI indices derived from each protocol, before and after ComBat harmonization.

### 3.2 Age Effects in Cognitively Normal Participants

To further investigate the effects of ComBat and statistical dMRI harmonization on important biologically driven dMRI differences, age associations were assessed in CN participants. Age effects were similar across methods and dMRI measures, with FA results showing the most variability. For AxD, 25 out of the 28 ROIs were consistently significant while the GCC and BCC were not significant across any of the 16 approaches; Fx associations were only significant in the Raw(Site) regression. For RD, all 28 ROIs were significant when harmonizing with any of the three ComBat variations, irrespective of the statistical model. However for ‘raw’ RD, all ROIs but the Fx were significant when using Raw(Site) or Raw(Protocol|Site), 23 ROIs were significant using Raw(Protocol), and 21 ROIs using Raw(None). For ‘raw’ and ComBat-harmonized MD data, all 28 ROIs were significant using either *site* or *protocol*|*site* as a grouping variable. With no random effect or with *protocol* as the grouping variable, only the BCC and GCC were not significant; again, the Fx was only significant in Raw(Site) or Raw(Protocol|Site) regressions. The same 21 out of 28 ROIs were significantly associated with age across all ComBat-harmonized FA metrics, regardless of statistical model; the SLF was additionally significant in the CovBat(Protocol) and CovBat(None) regressions. Only 18 FA ROIs were significant using Raw(None); the GCC, BCC, and Fx/ST were no longer significant. Consistently across tests, the most significant associations were found with SS AxD, ALIC RD, PLIC MD and ALIC FA. **Figure 5** illustrates the overall stability of associations across harmonization permutations by plotting the regional effects (*t*-values) of age in CNs on MD and FA in the cingulum (hippocampus) (CGH), fornix *(crus) / stria terminalis* (Fx/ST) and *genu* of corpus callosum (GCC) which showed the greatest effect sizes in prior ADNI3 analyses.^16^ Effect sizes were slightly boosted by using either ComBat or its variations, and by adjusting for *site* or *protocol*|*site* in the statistical model.

**Figure 5.**
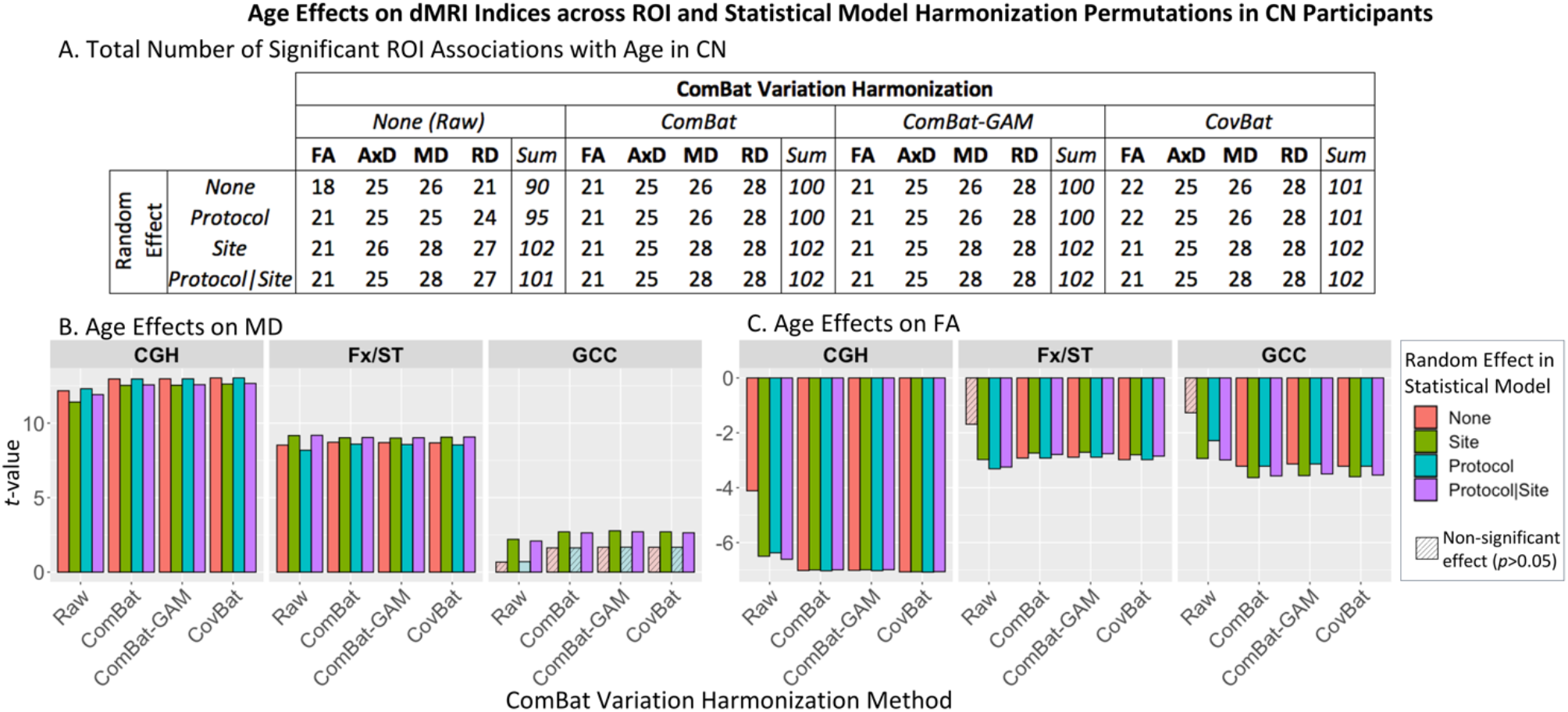
Age effects on dMRI indices across ROI and statistical model harmonization permutations in CN participants. (A) Total number of ROIs passing the FDR threshold (*q*=0.05). (B) Regional effects (*t*-values) of age, while covarying for sex and age*sex, on MD and FA in the CGH, Fx/ST and GCC, testing different combinations of ROI-level harmonization methods and random effects in the statistical model.

For each of the ComBat(Site), Raw(Protocol|Site) and Raw(None) regressions, paired t-tests were used to identify differences between ROI *t*-values resulting from associations between dMRI indices and age in CN participants. When comparing *t*-values from ComBat(Site) ROI regressions and Raw(Protoco|Site) regressions, only FA *t*-values were significantly different (*p*=6.32×10^−5^); the diffusivity metrics were not. ComBat(Site) AxD (*p*=0.04), FA (*p*=9.25×10^−11^) and RD (*p*=0.001) ROI *t*-values were also different from Raw(None). Finally, we also found FA (*p*=9.31×10^−8^), MD (*p*=0.009) and RD (*p*=2.61×10^−5^) differences between Raw(Protocol|Site) and Raw(None) regressions.

### 3.3 Assessing Effects of Age, Clinical Measures and Amyloid Status on dMRI Indices

As there were minimal differences between the 12 ComBat and statistical harmonization permutations, absolute regional effects of age, CDR-sob scores and Aβ-status are plotted in **Figure 6** only for Raw(Protocol|Site) and ComBat(Site) regressions. Overall, significance and effect sizes of associations were similar across harmonization approaches. Age effects were widespread, while CDR-sob effects were more limited, and Aβ-status associations were the weakest overall. The TAP AxD (‘raw’: *d*=1.15, ComBat: *d*=1.05) and MD (‘raw’: *d*=1.18, ComBat: *d*=1.15) and ALIC RD (raw: *d*=1.16, ComBat: *d*=1.15) and FA (raw: *d*=-1.10, ComBat: *d*=-1.07) were most strongly associated with age (**Figure 6A**). Notably, the Fx/ST stood out as the most strongly associated with CDR-sob (**Figure 6B**) for all three diffusivity metrics (AxD ‘raw’: *d*=0.8, ComBat: *d*=0.74; MD ‘raw’: *d=*1.08, ComBat: *d*=1.06; RD ‘raw’: *d=*1.11, ComBat: *d*=1.11), consistent with its role as a major commissure linking the hippocampus with the rest of the brain; the UNC was most strongly associated with CDR-sob for FA (‘raw’: *d*=0.84, ComBat: *d*=0.83). Aβ-positivity (**Figure 6C**) did not show many significant effects on dMRI indices, with the exception of MCP with the ‘raw’ FA, ICP, ML and CST with ‘raw’ AxD, and CGC and PTR with ComBat-harmonized AxD. Overall, for each harmonization approach, the largest effect sizes of Aβ-status on dMRI indices were yielded in the PTR (AxD raw: *D*=0.27, ComBat: *D*=0.29; MD raw: D=0.22, ComBat: *D*=0.25), CP (RD ComBat: *D*=0.21), and MCP (RD ‘raw’ *D*=-0.22; FA raw: *D*=0.31, ComBat: *D*=0.26).

**Figure 6.**
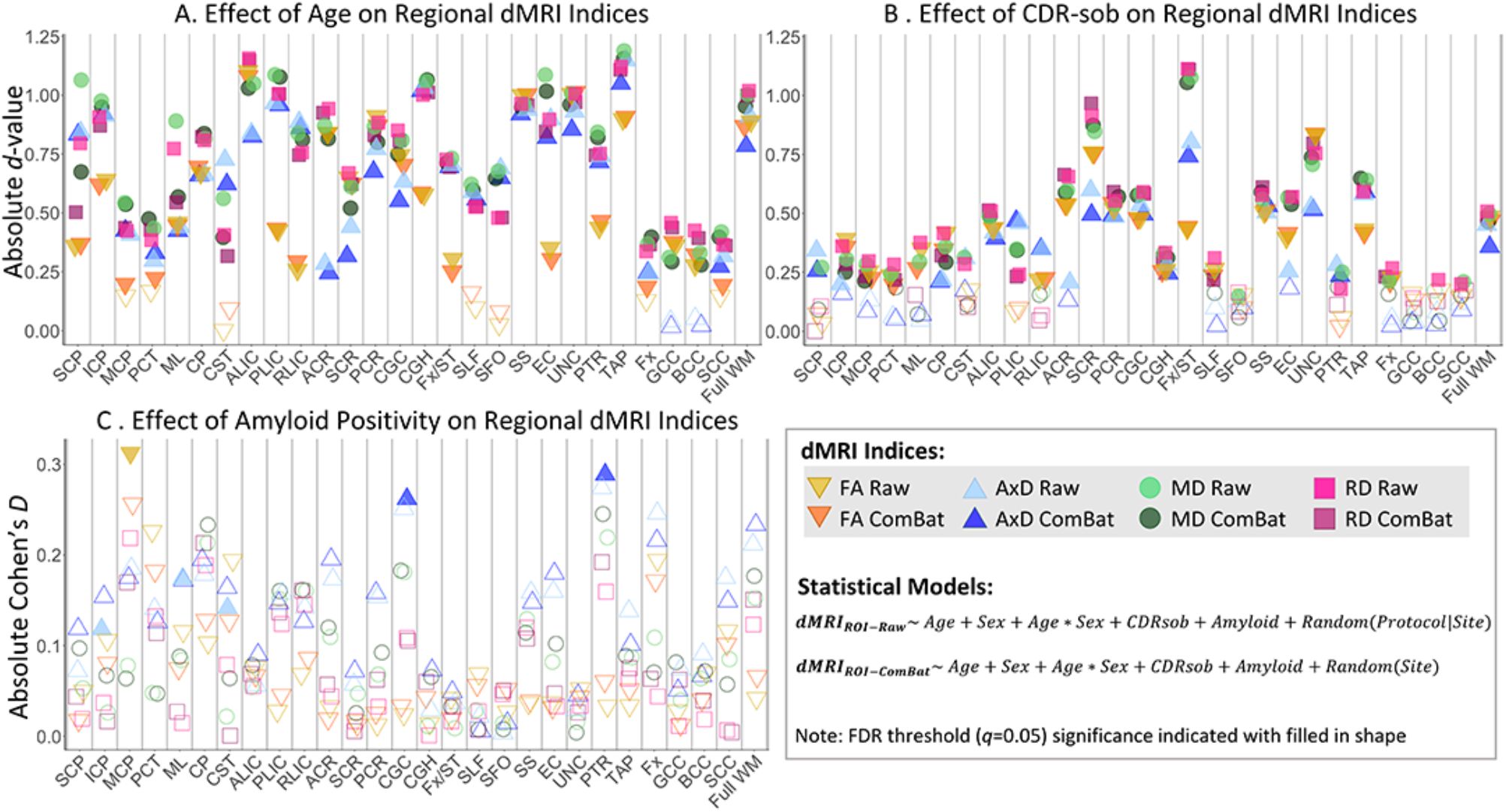
Absolute value of effect sizes (*d*-values) for regional dMRI associations with (A) age and (B) CDR-sob, in both ‘raw’ statistically-harmonized data or ComBat and statistically-harmonized data. Absolute Cohen’s *D* effect sizes for regional dMRI associations with (C) amyloid positivity are also plotted. ROIs surviving the FDR significance threshold (*q*=0.05) are indicated by a filled shape. While absolute values are plotted, for each panel the following ROIs had negative associations: all FA associations with age (A); GCC and ML AxD, and all but SCP, PLIC and RLIC FA associations with CDR-sob (B); and MCP, PCT, SLF, UNC MD, and MCP, PCT, Fx and UNC RD associations with amyloid positivity (C). We also observed negative associations with amyloid positivity in the ComBat-harmonized SCC RD and ‘raw’ CST, ML ICP, SLF RD, and in the ‘raw’ SLF AxD and ComBat-harmonized SFO AxD. Finally, we saw negative associations with CP, ALIC, PLIC, RLIC, PTR, SS, CGH, Fx/ST, SFO, TAP, and full WM in both ‘raw’ and ComBat-harmonized FA, in addition to the ComBat-harmonized GCC FA.

## 4. DISCUSSION

Overall, we found no significant differences among the harmonization methods tested. Age effects were slightly boosted when applying either ComBat or its variations, and/or by adjusting for *site* or *protocol*|*site* in the statistical model. Demographic and clinical heterogeneity in the raw data is highly reduced in ADNI due to the strict inclusion and exclusion criteria (e.g., exclusion of individuals with substantial vascular disease). We detected differences in the distributions of measures derived from data acquired across sites and scanner acquisition protocols, but the consistent overall design of ADNI may mean that random effects modeling of site is largely sufficient, without large gains from more sophisticated corrections. ComBat-GAM is likely to be more advantageous for multisite studies with a wider age range, or a difference in the age range across centers, where nonlinear effects of age are important. Although the current study had a wide age range (55.1-95.9 years), the average age was comparable across acquisition protocols and diagnostic groups. However, the number of participants in each diagnostic group varied for the different dMRI protocols, especially for the dementia group (see **Table 1** for example, S55 N=24 vs P33 N=3 diagnosed with dementia), making it more difficult to make direct comparisons of the effects of acquisition protocol and batch effects without having the same participants within each diagnostic group at each site.

Current ongoing work, addressing limitations seen in the present study, includes increasing our sample size as more data becomes available through ADNI, assessing cross-sectional harmonization methods at the voxel level, and testing harmonization methods for longitudinal dMRI data^60^ to compare method and rank dMRI metrics for their associations with key measures of AD biology and severity. Future work may detect greater benefit from more sophisticated harmonization methods, especially as more diverse cohorts beyond ADNI, are integrated. Choice of dMRI model may also influence harmonization performance.^27,61^ The effects of harmonization on multi-shell data is also being evaluated^62–65^ in an effort to model and investigate intra- and extracellular diffusion, neurite dispersion, and diffusion kurtosis with relation to aging, dementia and brain amyloid load.

Comparisons of harmonization methods are increasingly needed in the field of neuroimaging, and the development of these methods has proven highly valuable in the effort to assess heterogenous, multisite data. As mentioned above, ADNI’s participant recruitment is already relatively harmonized with strict inclusion and exclusion criteria, as it was designed to resemble a clinical trial population.^66^ ADNI’s fourth phase will aim to address this criticism and recruit participants that are more representative of the general population, and harmonization methods will play a crucial role in assessing future ADNI data. Other multisite studies without harmonized recruitment across centers – such as those conducted by the Enhancing NeuroImaging Genetics through Meta-Analysis (ENIGMA) and the Cohorts for Heart and Aging Research in Genomic Epidemiology (CHARGE) Consortia – will benefit from adding more diverse datasets with deeper phenotyping to better model confounds and understand selection biases. Pooling data from different centers, cities and countries has allowed for studies, especially those of genetics, to increase their power without the need to acquire more data.

## Data Availability

Not applicable.

## 5. ACKNOWLEDGEMENTS & DISCLOSURES

Data collection and sharing for ADNI was funded by National Institutes of Health Grant U01 AG024904 and the DOD (Department of Defense award number W81XWH-12-2-0012). This study was also supported by NIH grants U01AG068057, P41EB015922, and T32 T32AG058507. PMT and NJ received a research grant from Biogen, Inc. for research unrelated to this manuscript.

## REFERENCES

[1] World Health Organization (W., “Dementia,” <https://www.who.int/news-room/fact-sheets/detail/dementia> (5 July 2021).

[2] Jack, C. R., Bennett, D. A., Blennow, K., Carrillo, M. C., Dunn, B., Haeberlein, S. B., Holtzman, D. M., Jagust, W., Jessen, F., Karlawish, J., Liu, E., Molinuevo, J. L., Montine, T., Phelps, C., Rankin, K. P., Rowe, C. C., Scheltens, P., Siemers, E., Snyder, H. M., et al., “NIA-AA Research Framework: Toward a biological definition of Alzheimer’s disease,” Alzheimer’s Dement. 14(4), 535–562 (2018).

[3] Jack, C. R. J., Bennett, D. A., Blennow, K., Carrillo, M. C., Feldman, H. H., Frisoni, G. B., Hampel, H., Jagust, W. J., Johnson, K. A., Knopman, D. S., Petersen, R. C., Scheltens, P., Sperling, R. A. and Dubois, B., “A/T/N: An unbiased descriptive classification scheme for Alzheimer disease biomarkers,” Neurology 87(5), 539–547 (2016).

[4] Sullivan, E. V and Pfefferbaum, A., “Diffusion tensor imaging and aging,” Neurosci. Biobehav. Rev. 30(6), 749–761 (2006).

[5] Zhang, H., Schneider, T., Wheeler-Kingshott, C. A. and Alexander, D. C., “NODDI: Practical in vivo neurite orientation dispersion and density imaging of the human brain,” Neuroimage 61(4), 1000–1016 (2012).

[6] Özarslan, E., Koay, C. G., Shepherd, T. M., Komlosh, M. E., İrfanoğlu, M. O., Pierpaoli, C. and Basser, P. J., “Mean apparent propagator (MAP) MRI: A novel diffusion imaging method for mapping tissue microstructure,” Neuroimage 78, 16–32 (2013).

[7] Jensen, J. H., Helpern, J. A., Ramani, A., Lu, H. and Kaczynski, K., “Diffusional kurtosis imaging: The quantification of non-gaussian water diffusion by means of magnetic resonance imaging,” Magn. Reson. Med. 53(6), 1432–1440 (2005).

[8] Weiner, M. W., Veitch, D. P., Aisen, P. S., Beckett, L. A., Cairns, N. J., Green, R. C., Harvey, D., Jack, C. R. J., Jagust, W., Morris, J. C., Petersen, R. C., Salazar, J., Saykin, A. J., Shaw, L. M., Toga, A. W. and Trojanowski, J. Q., “The Alzheimer’s Disease Neuroimaging Initiative 3: Continued innovation for clinical trial improvement,” Alzheimers. Dement. 13(5), 561–571 (2017).

[9] Button, K. S., Ioannidis, J. P. A., Mokrysz, C., Nosek, B. A., Flint, J., Robinson, E. S. J. and Munafò, M. R., “Power failure: why small sample size undermines the reliability of neuroscience,” Nat. Rev. Neurosci. 14(5), 365–376 (2013).

[10] Thompson, P. M., Jahanshad, N., Ching, C. R. K., Salminen, L. E., Thomopoulos, S. I., Bright, J., Baune, B. T., Bertolín, S., Bralten, J., Bruin, W. B., Bülow, R., Chen, J., Chye, Y., Dannlowski, U., de Kovel, C. G. F., Donohoe, G., Eyler, L. T., Faraone, S. V, Favre, P., et al., “ENIGMA and global neuroscience: A decade of large-scale studies of the brain in health and disease across more than 40 countries,” Transl. Psychiatry 10(1), 100 (2020).

[11] Weiner, M. W., Veitch, D. P., Aisen, P. S., Beckett, L. A., Cairns, N. J., Green, R. C., Harvey, D., Jack, C. R. J., Jagust, W., Morris, J. C., Petersen, R. C., Saykin, A. J., Shaw, L. M., Toga, A. W. and Trojanowski, J. Q., “Recent publications from the Alzheimer’s Disease Neuroimaging Initiative: Reviewing progress toward improved AD clinical trials,” Alzheimers. Dement. 13(4), e1–e85 (2017).

[12] Gunter, J., Thostenson, K., Borowski, B., Reid, R., Arani, A., Bernstein, M., Fox, N., Thomas, D., Decarli, C., Tosun, D., Thompson, P., Weiner, M. and Jack Jr., C., “ADNI-3 MRI Protocol” (2017).

[13] Jovicich, J., Barkhof, F., Babiloni, C., Herholz, K., Mulert, C., van Berckel, B. N. M. and Frisoni, G. B., “Harmonization of neuroimaging biomarkers for neurodegenerative diseases: A survey in the imaging community of perceived barriers and suggested actions,” Alzheimer’s Dement. (Amsterdam, Netherlands) 11, 69–73 (2019).

[14] Nir, T. M., Jahanshad, N., Villalon-Reina, J. E., Toga, A. W., Jack, C. R., Weiner, M. W. and Thompson, P. M., “Effectiveness of regional DTI measures in distinguishing Alzheimer’s disease, MCI, and normal aging,” NeuroImage. Clin. 3, 180–195 (2013).

[15] Nir, T. M., Jahanshad, N., Villalon-Reina, J. E., Isaev, D., Zavaliangos-Petropulu, A., Zhan, L., Leow, A. D., Jack, C. R. J., Weiner, M. W. and Thompson, P. M., “Fractional anisotropy derived from the diffusion tensor distribution function boosts power to detect Alzheimer’s disease deficits,” Magn. Reson. Med. 78(6), 2322–2333 (2017).

[16] Zavaliangos-Petropulu, A., Nir, T. M., Thomopoulos, S. I., Reid, R. I., Bernstein, M. A., Borowski, B., Jack Jr., C. R., Weiner, M. W., Jahanshad, N. and Thompson, P. M., “Diffusion MRI Indices and Their Relation to Cognitive Impairment in Brain Aging: The Updated Multi-protocol Approach in ADNI3,” Front. Neuroinform. 13, 2 (2019).

[17] Vollmar, C., O’Muircheartaigh, J., Barker, G. J., Symms, M. R., Thompson, P., Kumari, V., Duncan, J. S., Richardson, M. P. and Koepp, M. J., “Identical, but not the same: Intra-site and inter-site reproducibility of fractional anisotropy measures on two 3.0T scanners,” Neuroimage 51(4), 1384–1394 (2010).

[18] Fox, R. J., Sakaie, K., Lee, J.-C., Debbins, J. P., Liu, Y., Arnold, D. L., Melhem, E. R., Smith, C. H., Philips, M. D., Lowe, M. and Fisher, E., “A validation study of multicenter diffusion tensor imaging: reliability of fractional anisotropy and diffusivity values,” AJNR. Am. J. Neuroradiol. 33(4), 695–700 (2012).

[19] Cannon, T. D., Sun, F., McEwen, S. J., Papademetris, X., He, G., van Erp, T. G. M., Jacobson, A., Bearden, C. E., Walker, E., Hu, X., Zhou, L., Seidman, L. J., Thermenos, H. W., Cornblatt, B., Olvet, D. M., Perkins, D., Belger, A., Cadenhead, K., Tsuang, M., et al., “Reliability of neuroanatomical measurements in a multisite longitudinal study of youth at risk for psychosis,” Hum. Brain Mapp. 35(5), 2424–2434 (2014).

[20] Zhu, A., Moyer, D., Nir, T., Thompson, P. and Jahanshad, N., “Challenges and Opportunities in dMRI Data Harmonization,” [Computational Diffusion MRI. MICCAI 2019. Mathematics and Visualization.], E. Bonet-Carne, F. Grussu, L. Ning, F. Sepehrband, and C. Tax, Eds., Spinger, Cham (2019).

[21] Moyer, D., Gao, S., Brekelmans, R., Galstyan, A. and Ver Steeg, G., “Invariant representations without adversarial training,” Adv. Neural Inf. Process. Syst. Montr. Canada Curran Assoc. Inc., 9102–9111 (2018).

[22] Moyer, D., Ver Steeg, G., Tax, C. M. W. and Thompson, P. M., “Scanner invariant representations for diffusion MRI harmonization,” Magn. Reson. Med. 84(4), 2174–2189 (2020).

[23] Dinsdale, N. K., Jenkinson, M. and Namburete, A. I. L., “Deep learning-based unlearning of dataset bias for MRI harmonisation and confound removal,” Neuroimage 228, 117689 (2021).

[24] Liu, M., Maiti, P., Thomopoulos, S. I., Zhu, A., Chai, Y., Kim, H. and Jahanshad, N., “Style Transfer Using Generative Adversarial Networks for Multi-Site {MRI} Harmonization,” bioRxiv (2021).

[25] Zuo, L., Dewey, B. E., Carass, A., Liu, Y., He, Y., Calabresi, P. A. and Prince, J. L., “Information-based Disentangled Representation Learning for Unsupervised MR Harmonization” (2021).

[26] Johnson, W. E., Li, C. and Rabinovic, A., “Adjusting batch effects in microarray expression data using empirical Bayes methods,” Biostatistics 8(1), 118–127 (2007).

[27] Fortin, J.-P., Parker, D., Tunç, B., Watanabe, T., Elliott, M. A., Ruparel, K., Roalf, D. R., Satterthwaite, T. D., Gur, R. C., Gur, R. E., Schultz, R. T., Verma, R. and Shinohara, R. T., “Harmonization of multi-site diffusion tensor imaging data,” Neuroimage 161, 149–170 (2017).

[28] Pomponio, R., Erus, G., Habes, M., Doshi, J., Srinivasan, D., Mamourian, E., Bashyam, V., Nasrallah, I. M., Satterthwaite, T. D., Fan, Y., Launer, L. J., Masters, C. L., Maruff, P., Zhuo, C., Völzke, H., Johnson, S. C., Fripp, J., Koutsouleris, N., Wolf, D. H., et al., “Harmonization of large MRI datasets for the analysis of brain imaging patterns throughout the lifespan,” Neuroimage 208, 116450 (2020).

[29] Chen, A. A., Beer, J. C., Tustison, N. J., Cook, P. A., Shinohara, R. T., Shou, H. and Initiative, the A. D. N., “Removal of Scanner Effects in Covariance Improves Multivariate Pattern Analysis in Neuroimaging Data,” bioRxiv, 858415 (2020).

[30] Kia, S. M., Huijsdens, H., Dinga, R., Wolfers, T., Mennes, M., Andreassen, O. A., Westlye, L. T., Beckmann, C. F. and Marquand, A. F., “Hierarchical Bayesian Regression for Multi-Site Normative Modeling of Neuroimaging Data,” arXiv (2020).

[31] Bayer, J. M. M., Dinga, R., Kia, S. M., Kottaram, A. R., Wolfers, T., Lv, J., Zalesky, A., Schmaal, L. and Marquand, A., “Accommodating site variation in neuroimaging data using hierarchical and Bayesian models,” bioRxiv, 2021.02.09.430363 (2021).

[32] Berg, L., “Clinical Dementia Rating (CDR),” Psychopharmacol. Bull. 24(4), 637–639 (1988).

[33] Landau, S. M., Thomas, B. A., Thurfjell, L., Schmidt, M., Margolin, R., Mintun, M., Pontecorvo, M., Baker, S. L. and Jagust, W. J., “Amyloid PET imaging in Alzheimer’s disease: a comparison of three radiotracers,” Eur. J. Nucl. Med. Mol. Imaging 41(7), 1398–1407 (2014).

[34] Landau, S. M., Breault, C., Joshi, A. D., Pontecorvo, M., Mathis, C. A., Jagust, W. J. and Mintun, M. A., “Amyloid-β imaging with Pittsburgh compound B and florbetapir: comparing radiotracers and quantification methods,” J. Nucl. Med. 54(1), 70–77 (2013).

[35] Landau, S. M., Mintun, M. A., Joshi, A. D., Koeppe, R. A., Petersen, R. C., Aisen, P. S., Weiner, M. W. and Jagust, W. J., “Amyloid deposition, hypometabolism, and longitudinal cognitive decline,” Ann. Neurol. 72(4), 578–586 (2012).

[36] Landau, S. M., Lu, M., Joshi, A. D., Pontecorvo, M., Mintun, M. A., Trojanowski, J. Q., Shaw, L. M. and Jagust, W. J., “Comparing positron emission tomography imaging and cerebrospinal fluid measurements of β-amyloid,” Ann. Neurol. 74(6), 826–836 (2013).

[37] Thomopoulos, S. I., Nir, T. M., Villalon-Reina, J. E., Haddad, E., Jahanshad, N., Reid, R., Bernstein, M. A., Borowski, B., Clifford R. Jack, J., Weiner, M. W. and Thompson, P. M., “Detection of Aging Effect on White Matter Microstructure: A Comparison of Diffusion MRI Preprocessing Pipelines,” SFN 2019 (2019).

[38] Garyfallidis, E., Brett, M., Amirbekian, B., Rokem, A., van der Walt, S., Descoteaux, M. and Nimmo-Smith, I., “Dipy, a library for the analysis of diffusion MRI data,” Front. Neuroinform. 8, 8 (2014).

[39] Manjón, J. V, Coupé, P., Concha, L., Buades, A., Collins, D. L. and Robles, M., “Diffusion weighted image denoising using overcomplete local PCA,” PLoS One 8(9), e73021 (2013).

[40] Veraart, J., Fieremans, E. and Novikov, D. S., “Diffusion MRI noise mapping using random matrix theory,” Magn. Reson. Med. 76(5), 1582–1593 (2016).

[41] Veraart, J., Novikov, D. S., Christiaens, D., Ades-Aron, B., Sijbers, J. and Fieremans, E., “Denoising of diffusion MRI using random matrix theory,” Neuroimage 142, 394–406 (2016).

[42] Tournier, J.-D., Smith, R., Raffelt, D., Tabbara, R., Dhollander, T., Pietsch, M., Christiaens, D., Jeurissen, B., Yeh, C.-H. and Connelly, A., “MRtrix3: A fast, flexible and open software framework for medical image processing and visualisation,” Neuroimage 202, 116137 (2019).

[43] Kellner, E., Dhital, B., Kiselev, V. G. and Reisert, M., “Gibbs-ringing artifact removal based on local subvoxel-shifts,” Magn. Reson. Med. 76(5), 1574–1581 (2016).

[44] Woolrich, M. W., Jbabdi, S., Patenaude, B., Chappell, M., Makni, S., Behrens, T., Beckmann, C., Jenkinson, M. and Smith, S. M., “Bayesian analysis of neuroimaging data in FSL,” Neuroimage 45(1 Suppl), S173–86 (2009).

[45] Smith, S. M., Jenkinson, M., Woolrich, M. W., Beckmann, C. F., Behrens, T. E. J., Johansen-Berg, H., Bannister, P. R., De Luca, M., Drobnjak, I., Flitney, D. E., Niazy, R. K., Saunders, J., Vickers, J., Zhang, Y., De Stefano, N., Brady, J. M. and Matthews, P. M., “Advances in functional and structural MR image analysis and implementation as FSL,” Neuroimage 23 Suppl 1, S208–19 (2004).

[46] Jenkinson, M., Beckmann, C. F., Behrens, T. E. J., Woolrich, M. W. and Smith, S. M., “FSL,” Neuroimage 62(2), 782–790 (2012).

[47] Andersson, J. L. R. and Sotiropoulos, S. N., “An integrated approach to correction for off-resonance effects and subject movement in diffusion MR imaging,” Neuroimage 125, 1063–1078 (2016).

[48] Andersson, J. L. R., Graham, M. S., Zsoldos, E. and Sotiropoulos, S. N., “Incorporating outlier detection and replacement into a non-parametric framework for movement and distortion correction of diffusion MR images,” Neuroimage 141, 556–572 (2016).

[49] Andersson, J. L. R., Graham, M. S., Drobnjak, I., Zhang, H., Filippini, N. and Bastiani, M., “Towards a comprehensive framework for movement and distortion correction of diffusion MR images: Within volume movement,” Neuroimage 152, 450–466 (2017).

[50] Isensee, F., Schell, M., Pflueger, I., Brugnara, G., Bonekamp, D., Neuberger, U., Wick, A., Schlemmer, H.-P., Heiland, S., Wick, W., Bendszus, M., Maier-Hein, K. H. and Kickingereder, P., “Automated brain extraction of multisequence MRI using artificial neural networks,” Hum. Brain Mapp. 40(17), 4952–4964 (2019).

[51] Fischl, B., van der Kouwe, A., Destrieux, C., Halgren, E., Ségonne, F., Salat, D. H., Busa, E., Seidman, L. J., Goldstein, J., Kennedy, D., Caviness, V., Makris, N., Rosen, B. and Dale, A. M., “Automatically parcellating the human cerebral cortex,” Cereb. Cortex 14(1), 11–22 (2004).

[52] Greve, D. N. and Fischl, B., “Accurate and robust brain image alignment using boundary-based registration,” Neuroimage 48(1), 63–72 (2009).

[53] Avants, B. B., Epstein, C. L., Grossman, M. and Gee, J. C., “Symmetric diffeomorphic image registration with cross-correlation: evaluating automated labeling of elderly and neurodegenerative brain,” Med. Image Anal. 12(1), 26–41 (2008).

[54] Avants, B. B., Tustison, N. J., Song, G., Cook, P. A., Klein, A. and Gee, J. C., “A reproducible evaluation of ANTs similarity metric performance in brain image registration,” Neuroimage 54(3), 2033–2044 (2011).

[55] Mori, S., Oishi, K., Jiang, H., Jiang, L., Li, X., Akhter, K., Hua, K., Faria, A. V, Mahmood, A., Woods, R., Toga, A. W., Pike, G. B., Neto, P. R., Evans, A., Zhang, J., Huang, H., Miller, M. I., van Zijl, P. and Mazziotta, J., “Stereotaxic white matter atlas based on diffusion tensor imaging in an ICBM template,” Neuroimage 40(2), 570–582 (2008).

[56] Basser, P. J., Mattiello, J. and LeBihan, D., “MR diffusion tensor spectroscopy and imaging,” Biophys. J. 66(1), 259–267 (1994).

[57] Wilcox, R. R., [Introduction to Robust Estimation and Hypothesis Testing, 3rd ed.], Academic Press, San Diego, CA (2012).

[58] Mair, P. and Wilcox, R., “Robust statistical methods in R using the WRS2 package,” Behav. Res. Methods 52(2), 464–488 (2020).

[59] Benjamini, Y. and Hochberg, Y., “Controlling the False Discovery Rate: A Practical and Powerful Approach to Multiple Testing,” J. R. Stat. Soc. Ser. B 57(1), 289–300 (1995).

[60] Beer, J. C., Tustison, N. J., Cook, P. A., Davatzikos, C., Sheline, Y. I., Shinohara, R. T. and Linn, K. A., “Longitudinal ComBat: A method for harmonizing longitudinal multi-scanner imaging data,” Neuroimage 220, 117129 (2020).

[61] Nir, T. M., Lam, H. Y., Ananworanich, J., Boban, J., Brew, B. J., Cysique, L., Fouche, J. P., Kuhn, T., Porges, E. S., Law, M., Paul, R. H., Thames, A., Woods, A. J., Valcour, V. G., Thompson, P. M., Cohen, R. A., Stein, D. J. and Jahanshad, N., “Effects of Diffusion MRI Model and Harmonization on the Consistency of Findings in an International Multi-cohort HIV Neuroimaging Study ,” Int. Work. Comput. Diffus. MRI, E. Bonet-Carne, F. Grussu, L. Ning, F. Sepehrband, and C. M. W. Tax, Eds., 203-215 BT-Computational Diffusion MRI (2019).

[62] Reina, J. E. V., Nir, T. M., Thomopoulos, S. I., Salminen, L., Jahanshad, N. and Thompson, P., “Evaluating NODDI-based biomarkers of Alzheimer’s disease: Neuroimaging/Optimal neuroimaging measures for early detection,” Alzheimer’s Dement. 16, e042297 (2020).

[63] Nir, T., Villalon-Reina, J., Thomopoulos, S., Zavaliangos-Petropulu, A., Reid, R., Bernstein, M., Borowski, B., Jack, Jr., C., Weiner, M., Jahanshad, N., Thompson, P. and for the Alzheimer’s Disease Neuroimaging Initiative., “Comparing NODDI Implementations for Evaluating Brain Microstructure with ADNI3 Diffusion MRI,” OHBM 2019 (2019).

[64] Villalon-Reina, J. E., Nir, T. M., Thomopoulos, S. I., Salminen, L. E., Jahanshad, N., Fick, R., Frigo, M., Deriche, R., Thompson, P. M. and for the Alzheimer’s Disease Neuroimaging Initiative (ADNI)., “Tracking microstructural biomarkers of Alzheimer’s disease via advanced multi-shell diffusion MRI scalar measures,” ISMRM 2020 (2020).

[65] Nir, T. M., Thomopoulos, S. I., Villalon-Reina, J. E., Zavaliangos-Petropulu, A., Dennis, E. L., Reid, R. I., Bernstein, M. A., Borowski, B., Jack, C. R., Weiner, M. W., Jahanshad, N. and Thompson, P. M., “Multi-Shell Diffusion MRI Measures of Brain Aging: A Preliminary Comparison From ADNI3,” 2019 IEEE 16th Int. Symp. Biomed. Imaging (ISBI 2019), 173–177, IEEE (2019).

[66] Petersen, R. C., Aisen, P. S., Beckett, L. A., Donohue, M. C., Gamst, A. C., Harvey, D. J., Jack, C. R., Jagust, W. J., Shaw, L. M., Toga, A. W., Trojanowski, J. Q. and Weiner, M. W., “Alzheimer’s Disease Neuroimaging Initiative (ADNI): Clinical characterization,” Neurology 74(3), 201–209 (2010).

